# Automated Extraction of Stroke Severity from Unstructured Electronic Health Records using Natural Language Processing

**DOI:** 10.1101/2024.03.08.24304011

**Authors:** Marta Fernandes, M. Brandon Westover, Aneesh B. Singhal, Sahar F. Zafar

## Abstract

**BACKGROUND:** Multi-center electronic health records (EHR) can support quality improvement initiatives and comparative effectiveness research in stroke care. However, limitations of EHR-based research include challenges in abstracting key clinical variables from non-structured data at scale. This is further compounded by missing data. Here we develop a natural language processing (NLP) model that automatically reads EHR notes to determine the NIH stroke scale (NIHSS) score of patients with acute stroke.

**METHODS:** The study included notes from acute stroke patients (>= 18 years) admitted to the Massachusetts General Hospital (MGH) (2015-2022). The MGH data were divided into training (70%) and hold-out test (30%) sets. A two-stage model was developed to predict the admission NIHSS. A linear model with the least absolute shrinkage and selection operator (LASSO) was trained within the training set. For notes in the test set where the NIHSS was documented, the scores were extracted using regular expressions (stage 1), for notes where NIHSS was not documented, LASSO was used for prediction (stage 2). The reference standard for NIHSS was obtained from Get With The Guidelines Stroke Registry. The two-stage model was tested on the hold-out test set and validated in the MIMIC-III dataset (Medical Information Mart for Intensive Care-MIMIC III 2001-2012) v1.4, using root mean squared error (RMSE) and Spearman correlation (SC).

**RESULTS:** We included 4,163 patients (MGH = 3,876; MIMIC = 287); average age of 69 [SD 15] years; 53% male, and 72% white. 90% patients had ischemic stroke and 10% hemorrhagic stroke. The two-stage model achieved a RMSE [95% CI] of 3.13 [2.86-3.41] (SC = 0.90 [0.88-0. 91]) in the MGH hold-out test set and 2.01 [1.58-2.38] (SC = 0.96 [0.94-0.97]) in the MIMIC validation set.

**CONCLUSIONS:** The automatic NLP-based model can enable large-scale stroke severity phenotyping from EHR and therefore support real-world quality improvement and comparative effectiveness studies in stroke.

## Introduction

Real world studies using multi-center electronic health records (EHR) can pave the way for understanding patterns and practice variation in stroke care that can support process improvement and treatment decisions [1]. EHR can be particularly useful for quality of care improving efforts [2,3], investigating and addressing disparities in health care [4,5], understanding gaps in health care delivery [6], and designing effective measures to improve patient outcomes [7,8]. However, limitations of EHR-based research include challenges in abstracting key clinical variables, such as stroke severity, from non-structured data at scale [9]. This is further compounded by missing data [10]. Accurately measuring stroke severity from EHR is critical for large-scale comparative effectiveness research and quality improvement [11,12].

The National Institutes of Health Stroke Scale (NIHSS) is the gold standard for measuring stroke severity in the clinical environment [9]. The Joint Commission requires the NIHSS to be performed on all patients that receive thrombolytics or undergo thrombectomy, and those presenting within 12 hours of symptom onset [13]. Unfortunately, NIHSS is not always documented in the EHR and when documented, it is frequently in clinical notes and not as structured data [9]. This creates limitations for performing population level research or conducting quality improvement, particularly in the case of acute ischemic stroke, patients not meeting the criteria for acute treatments or interventions [14], patients seen at smaller community or non-stroke centers [15–17] or admitted to non-Joint Commission accredited centers [17]. An additional limitation is that NIHSS is not always documented for hemorrhagic stroke patients. While scores including the Intracerebral Hemorrhage and Hunt and Hess scores are used for hemorrhagic stroke patients, the NIHSS can be used to predict mortality and correlates with hemorrhage volume [18,19], and serves as a common score across all stroke subtypes for population level research [20,21]. NIHSS can be abstracted from clinical notes by chart review, however this is labor intensive [22]. Prior studies have developed machine learning models using structured data, such as International Classification of Diseases (ICD) codes and Current Procedural Terminology (CPT) codes to measure stroke severity [23]. However, models only restricted to structured data, besides not considering missing data [10], leave out the extensive data available in unstructured notes that would allow for more accurate phenotyping. Existing natural language processing (NLP) models can only be applied to notes with documented NIHSS score or its subcomponents, and therefore not applicable for missing data, and have not been validated in other datasets [24]. The lack of standardization in stroke severity assessment, documentation and reporting in EHR databases precludes the advancement of population level stroke-related comparative effectiveness research [25,26].

Here we aimed to develop an NLP model that automatically reads EHR clinical notes to accurately predict the NIHSS score of patients presenting with acute stroke. We used regional EHR linked to the American Heart Association’s (AHA) Get With The Guidelines (GWTG) – Stroke Registry [27] as the gold standard dataset and validated the model on an external independent dataset. This model is intended to enable large-scale EHR stroke severity phenotyping even when the NIHSS or its subcomponents are not documented or recorded.

## Methods

### Study Population

We included adult patients (≥ 18 years old) with acute stroke (ischemic and hemorrhagic). The study was approved by the Mass General Brigham Institutional Review Board; a waiver of informed consent was obtained for this observational study. This study consists of retrospective data analysis and is reported in accordance with the STrengthening the Reporting of OBservational studies in Epidemiology (STROBE) statement [28].

### Datasets

Our stroke cohort was derived from two sources: 1) patients admitted with acute stroke at Massachusetts General Hospital (MGH) between March 2015 and December 2022 were identified through the AHA GWTG – Stroke Registry linked with EHR [27]; 2) stroke admissions from the publicly available de-identified database Medical Information Mart for Intensive Care III (MIMIC-III) v1.4 [29], which contains medical records for ICU admissions at Beth Israel Deaconess Medical Center (BIDMC) between 2001 and 2012. Patients with acute stroke in the MIMIC dataset were identified using the ICD coding system [24]. Further details on cohort derivation are provided in Supplementary Information.

### Clinical notes

The EHR data in our study comprised free text admission notes (MGH) and discharge summaries (MIMIC), which consist of semi-structured text written by physicians. The MGH notes were extracted for the first and second dates of admission, given our goal to measure and predict admission stroke severity. All available notes were extracted, including among others, any neurology notes, emergency department, assessment and plan, history and physical (H&P), operative, procedures, consults, discharge instructions, discharge summaries, hospital course, progress, transfer, nursing, physical therapy and occupational therapy notes. For model external validation, we used discharge summaries from the MIMIC dataset, since the NIHSS scores are mainly stored in this type of notes. The discharge summaries include the patients’ initial history and physical and admission clinical exams. The notes from each patient were preprocessed (see Table S.1.) and converted into a structured format consisting of binary variables. Each variable indicated the presence or absence of an n-gram (single word [unigram], or sequence of 2 [bigrams] or 3 [trigrams] words) in the notes (see Supplementary Information).

### Outcomes and gold standard

Our outcome was the initial (admission) total NIHSS score. For the MGH cohort, the gold standard scores were obtained from the AHA GWTG – Stroke Registry [27]. For the MIMIC cohort, gold standard scores were obtained by applying rule-based regular expressions (regexes) to the notes for extraction of the scores, followed by manual note review for expert validity by a neurologist/ neurointensivist (SFZ) (see Supplementary Information). After chart review, for MGH patients, notes with any discrepancy between the GWTG gold standard and the neurological exam documented in the note or the NIHSS, if also documented in the note, were removed. For MIMIC patients, any notes that did not have a documented NIHSS were removed.

### Statistical Analysis

First, we split the MGH data randomly into a training set (70%) and hold-out test set (30%), as in previous studies [30–32]. With the training data, we developed a linear regression model using the least absolute shrinkage and selection operator (LASSO) that utilized the text-based features from the notes to predict the patients’ NIHSS scores. We performed 100 iterations within the training data of five-fold cross validation to determine the best regularization parameter (see Supplementary Information). The relative importance of the variables was assessed by magnitude of the regression coefficients.

We then created a two-stage model, applied on the MGH hold-out test set and externally validated on the MIMIC validation set. (1) Stage 1: In the first stage notes were checked for the NIHSS and, if detected, the score was directly extracted. This stage used simple hard-coded regular expressions. (2) Stage 2: For notes where the NIHSS score was not detected/documented, we applied the LASSO model to estimate the NIHSS from information contained in the note.

To evaluate the model, we used the root mean squared error (RMSE) and Spearman correlation. We performed 1000 iterations of bootstrap random sampling with replacement to calculate 95% confidence intervals (CI).

We report overall results for the two-stage model on all notes in the MGH hold-out test set and MIMIC validation set (those that contain extractable NIHSS scores and those that do not). We also report results separately for stage 1 (for notes with extractable NIHSS) and stage 2 (for notes where the NIHSS scores were not documented and LASSO model was used to predict the score).

## Results

### Patients Characteristics

Our study cohort for analysis after inclusion and exclusion criteria (Figure 1) included a total of 4,163 patients (MGH n = 3,876; MIMIC n = 287) with an average age of 69 [SD 15] years, majority males (53%), White (72%) and presenting with ischemic stroke (90%). There were no observable differences in age, sex, race between train, test and external validation sets (Table 1). The median NIHSS was 5 vs 13 for the MGH vs MIMIC datasets. When looking at the distribution of NIHSS scores (Figure 2), we observe that the majority of patients in the MGH cohort had a score <5, while the MIMIC cohort displays a higher frequency of scores between 6 and 23. For approximately the entire MIMIC cohort (99%), the admissions were of emergency type, while for the MGH cohort the admissions included emergency (85%), urgent (14%) and elective (1%) types.

**Table 1.**
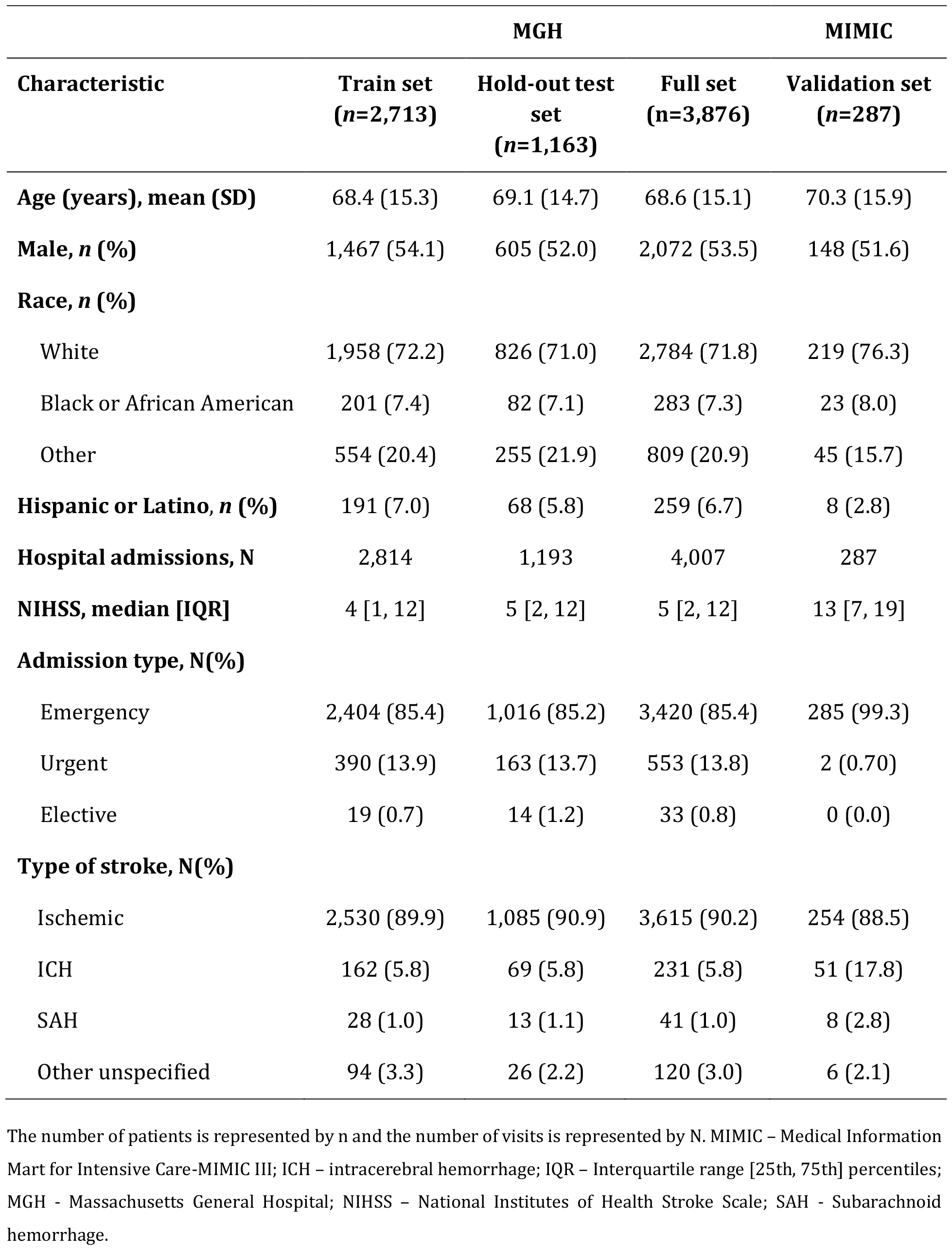
Characteristics of the study population stratified by train, test and external validation sets.

**Figure 1.**
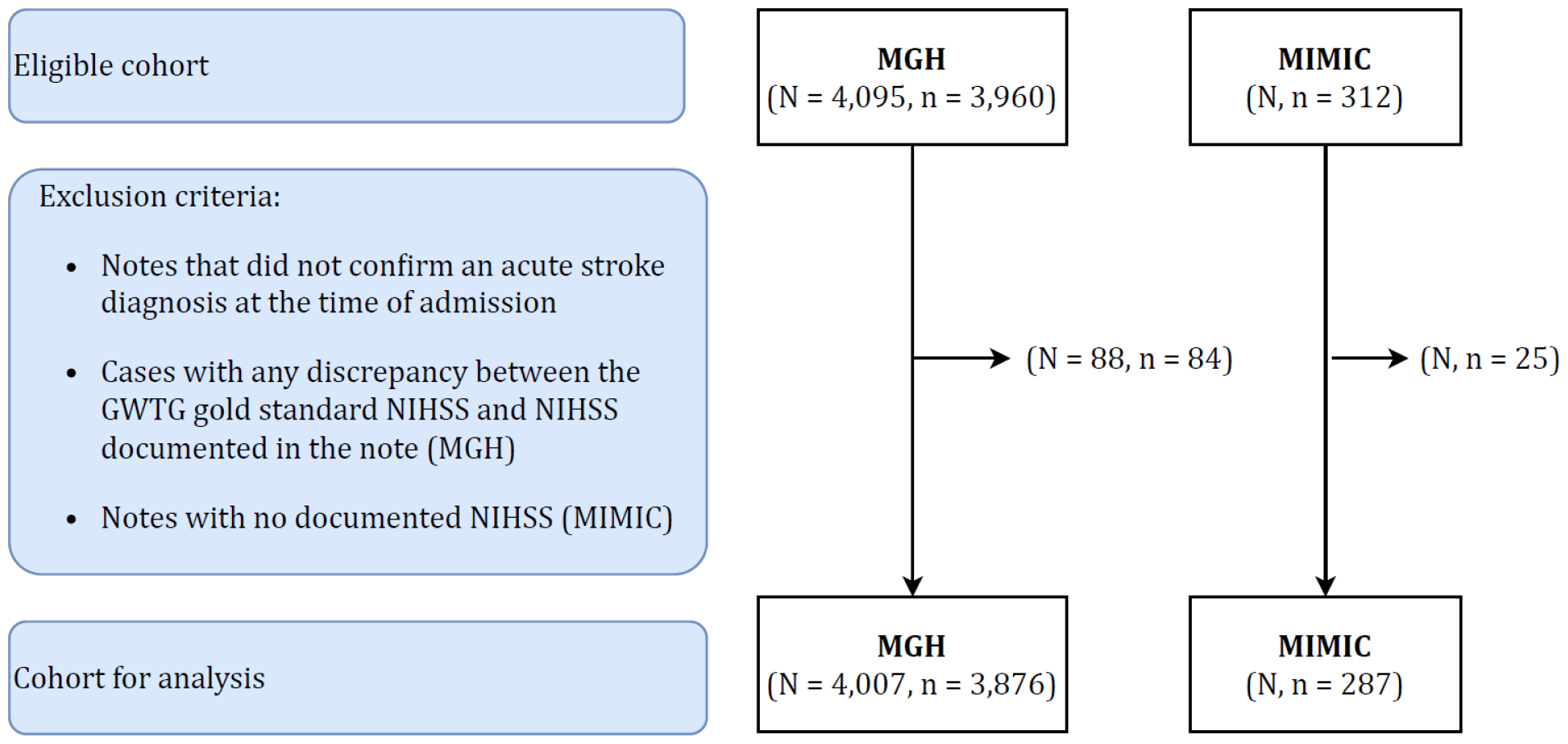
CONSORT (Consolidated Standards of Reporting Trials) chart. The number of patients is represented by ‘n’ and the number of visits by ‘N’. GWTG – Get With The Guidelines Stroke registry; MIMIC – Medical Information Mart for Intensive Care-MIMIC III; MGH - Massachusetts General Hospital; NIHSS – National Institutes of Health Stroke Scale.

**Figure 2.**
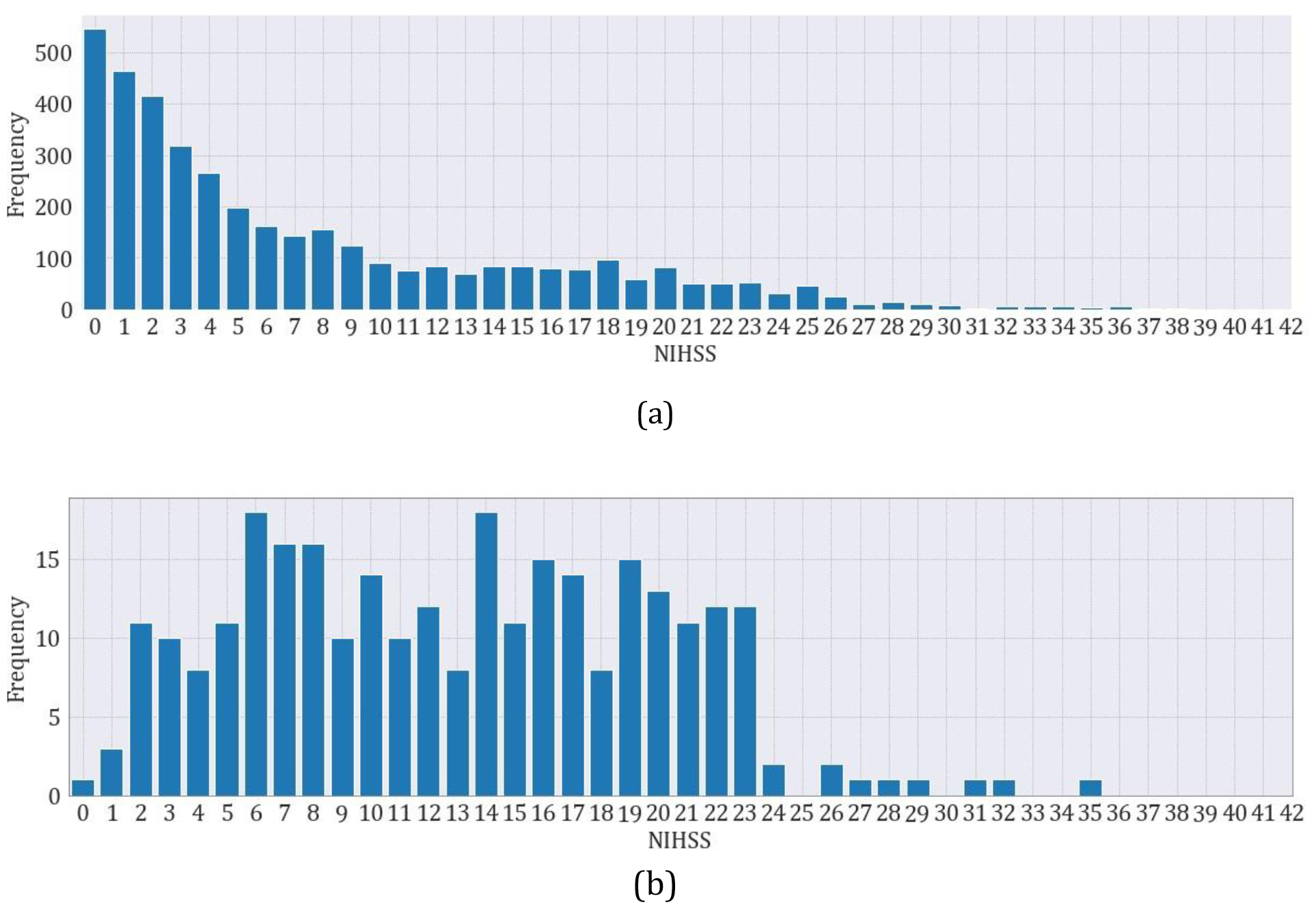
NIHSS score distribution for (a) MGH and (b) MIMIC cohorts. MIMIC – Medical Information Mart for Intensive Care-MIMIC III; MGH - Massachusetts General Hospital.

### Model Performance

The two-stage model was able to predict the NIHSS score with an RMSE of approximately 3 in the MGH hold-out test set and an RMSE of 2 in the MIMIC external validation set, as presented in table 2. For both, the Spearman correlation was at least 90%. The statistics regarding the number of documents with NIHSS utilized for the first stage of the model for both sets are presented in Supplementary Information, Table S.2. We observed that the number of visits with only one NIHSS documented was almost double for the external validation set (77%), compared to the hold-out test set (40%). MIMIC discharge summaries are more succinct narratives, when compared to MGH admission notes that tended to include narratives from more than one department and from multiple healthcare professionals.

**Table 2.** Performance of the two-stage model and each individual model stage, in 95% confidence intervals.

| **Cohort** | **Two-stage model** |  | **Stage 1 (regexes)** |  | **Stage 2 (LASSO)** |  |
| --- | --- | --- | --- | --- | --- | --- |
|  | **RMSE**<br>**[95% CI]** | **SC**<br>**[95% CI]** | **RMSE**<br>**[95% CI]** | **SC**<br>**[95% CI]** | **RMSE**<br>**[95% CI]** | **SC**<br>**[95% CI]** |
| MGH hold-out test set | 3.13<br>[2.86, 3.41] | 0.90<br>[0.88, 0.91] | 1.93<br>[1.49, 2.39] | 0.95<br>[0.93, 0.97] | 3.72<br>[3.38, 4.10] | 0.87<br>[0.86, 0.89] |
| MIMIC validation set | 2.01<br>[1.58, 2.38] | 0.96<br>[0.94, 0.97] | 0.61<br>[0.00, 1.01] | 1.00<br>[0.99, 1.00] | 4.00<br>[3.32, 4.70] | 0.81<br>[0.71, 0.89] |

MIMIC – Medical Information Mart for Intensive Care-MIMIC III; CI – confidence intervals; MGH – Massachusetts General Hospital; Regexes – regular expressions; RMSE – root mean squared error; SC – Spearman correlation.

### Error Analysis

The distribution of the two-stage model predicted vs target NIHSS is presented in Figure 3. We performed a manual review of the model errors to better understand the dispersion and identify the main types of misclassifications (see Supplementary Information, Table S.3).

**Figure 3.**
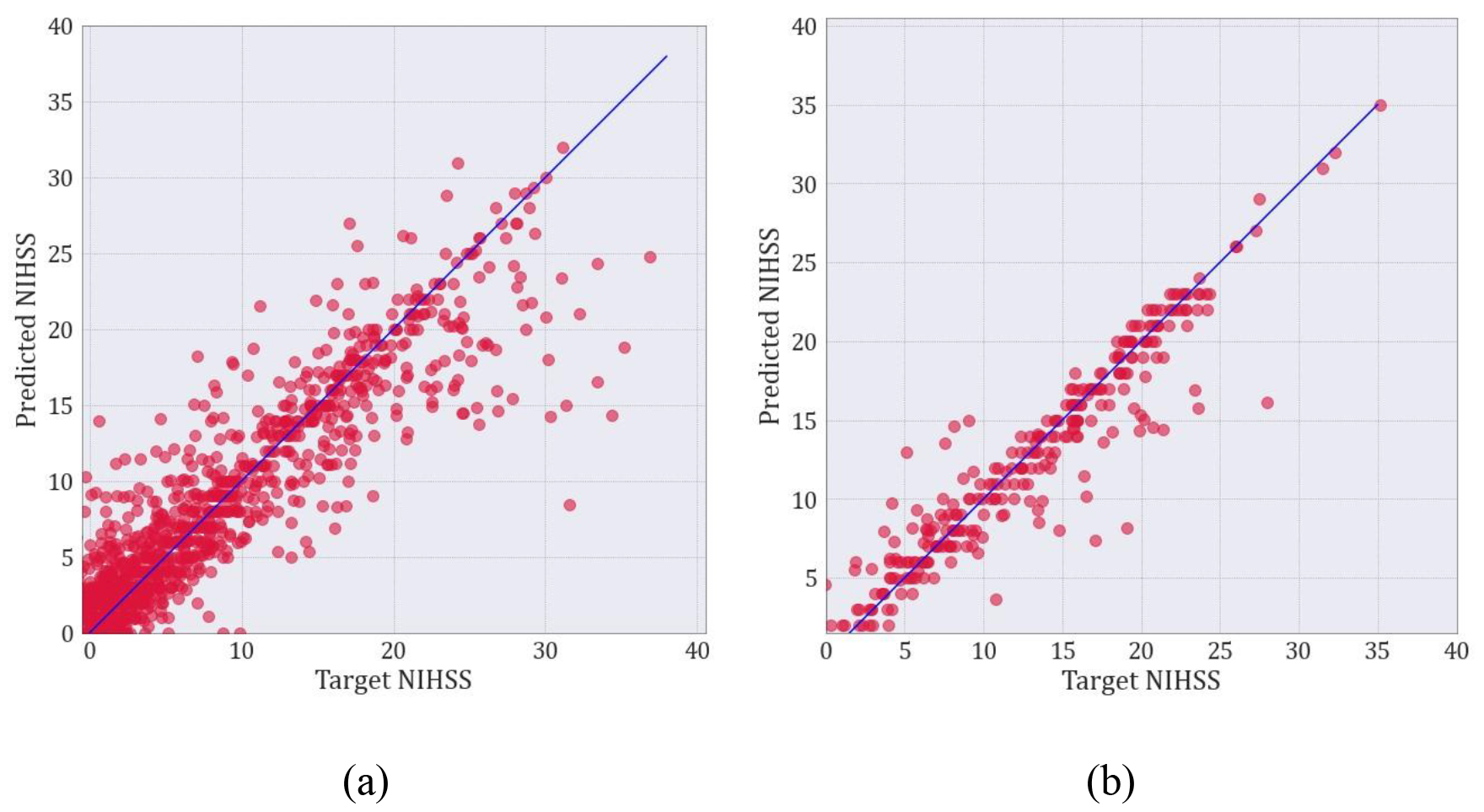
Two-stage model predicted versus target NIHSS scores in the (a) MGH hold-out test set and (b) MIMIC external validation set. MIMIC – Medical Information Mart for Intensive Care-MIMIC III; MGH – Massachusetts General Hospital.

First, we identified cases with fluctuating symptoms i.e. improvement or worsening of the neurological exam. These changes in exam occurred for example between teleconsultation/ outside hospital presentation and arrival to the stroke center ED, or between initial ED presentation and neurological consultation or admission to the ICU. In such cases the model either predicted one of the scores, an average or a score within the range documented. For example, in a case where the GWTG gold standard NIHSS was 30, and the note documented improvement in the NIHSS from 30 to 22, the model predicted an NIHSS of 21. This case yielded an absolute error of 9 compared with the first documented score, however, the model prediction score of 21 was very close to the improved score of 22.

A second type of error occurred when there was discrepancy between the sum of the NIHSS subcomponents and the documented NIHSS score. As an example, a note documented NIHSS as 20, while the individual subcomponents added to a total of 29. In this case the regexes captured “NIHSS 20” while the subcomponents added to 29 which was also the GWTG gold standard score.

We also identified cases where the note indicated a range of scores, but not the exact target score documented in the GWTG registry. An illustration of this scenario is a case where the NIHSS documented in the note was “15+/- 3”, the GTWG gold standard was 14, and the model prediction was 15.71. Even though in this example the error was relatively small, it demonstrated that in certain cases a model error up to 3 points might be acceptable.

Finally, we observed that our model performance decreased for NIHSS scores greater than 30. This is likely the result of smaller number of cases with NIHSS greater than 30, as depicted in Figure 2, and therefore not enough cases for model development and learning. Types of errors for greater scores included cases due to fluctuating symptoms, as already illustrated, and cases with fewer clinical details documented in the exam sections of the notes, with many of the notes documenting the patients were “comatose” or “unresponsive”, and fewer details on motor exam.

### Features Importance

The relative importance of the top 20 modeling variables is presented in Figure 4. The initial number of variables in the training vocabulary was 7,565, which was reduced to 347 by LASSO regularization (see Supplementary Information Table S.4). We observe that indication of lack of movement, not answering questions or following commands are all associated with higher stroke severity scores. Gaze deviation, use of urinary catheter, a middle cerebral artery (MCA) stroke, facial palsy and paralysis are also associated with higher scores. On the other hand, ‘drift’ contributed the most to lower prediction scores, being associated with lower scores for the NIHSS motor subcomponents (arms and legs). A level of consciousness of “alert and responsive”, as well as the word ‘deni’ (denies), meaning that the patient is responsive, also contributed to lower scores.

**Figure 4.**
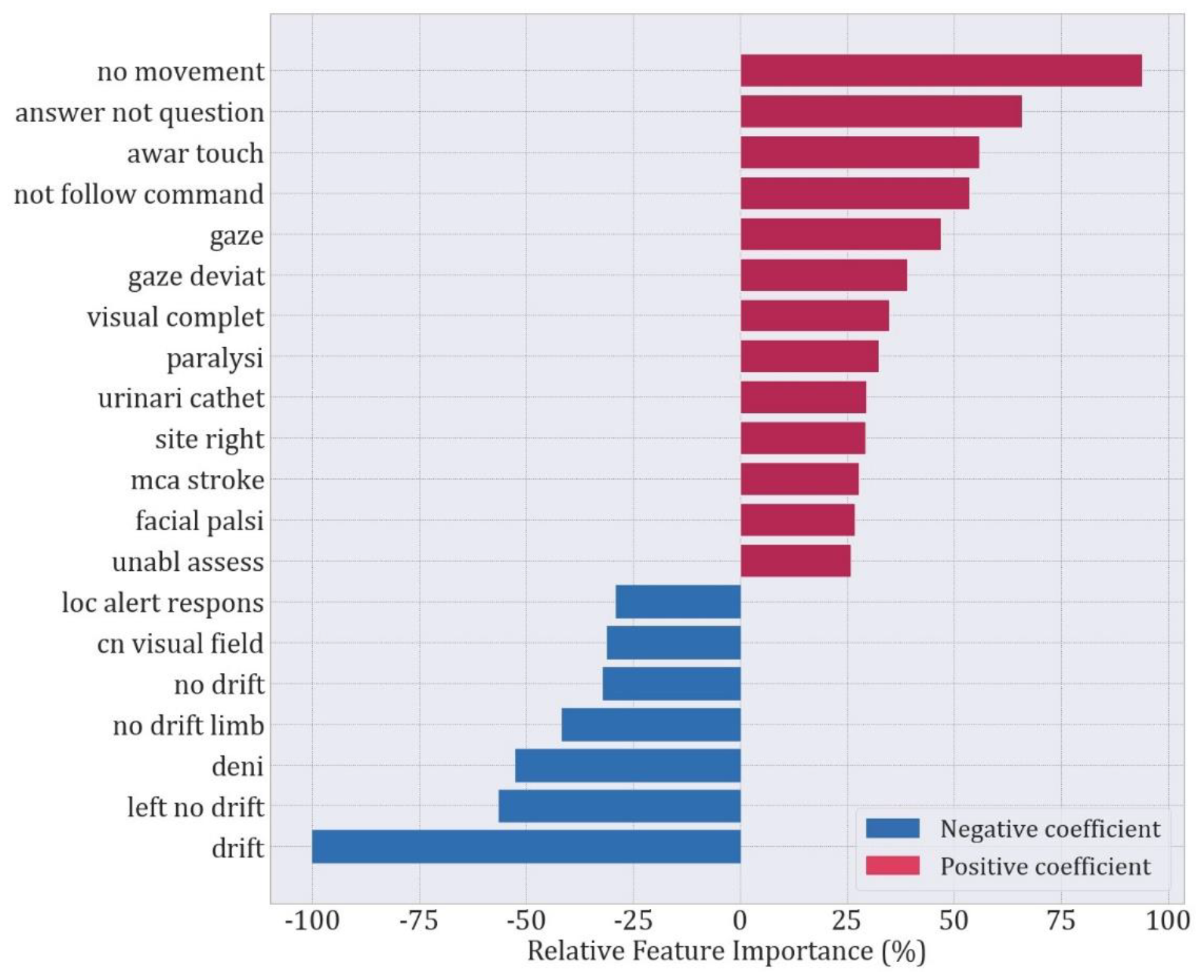
Relative feature importance given by the LASSO model coefficients for the top 20 features.

## Discussion

In this work we developed an NLP model that automatically “reads” EHR clinical notes to determine the NIHSS score of adult patients presenting with acute stroke. The model achieved good performance with an RMSE of approximately 3 and Spearman correlation of 90%. The modeling variables most indicative of higher scores were related to patients either being unresponsive, with MCA stroke, urinary catheter and other neurological deficits, such as facial palsy, paralysis or gaze deviation. The variables contributing the most to lower scores were related to the patients being alert and responsive, with lower scores in the motor subcomponents of the scale. Our findings suggest that automated EHR phenotyping of stroke is feasible in large and diverse cohorts.

A prior study [23] used administrative data including ICD and CPT codes, demographics, prescriptions/medications, hospital visit information, and comorbidities to predict the NIHSS. Using machine learning to assess stroke severity, the authors found that the main predictors included death within the same month as stroke occurrence, length of hospital stay following stroke occurrence, aphagia/dysphagia diagnosis, hemiplegia diagnosis, and whether a patient was discharged to home or self-care. Comparing the imputed NIHSS scores to the gold standard on the hold-out test set yielded a RMSE of 4.5 and Pearson correlation of 0.76. Based on the higher performance of our model, we hypothesize that using NLP and unstructured text notes can more accurately measure NIHSS, compared with using administrative data.

Another study [24], combined BERT-BiLSTM-CRF and a random forests model for the task of NIHSS item and score recognition, achieving an F1-score of 0.90, which outperformed their rule-based method (regexes) with F1-score of 0.81. The NIHSS item extraction showed best performance when using the rule-based method (regexes) with a precision of 1.00, recall of 0.95 and F1-score of 0.97. The study was, however, focused only on extraction of the scores from note when documented and therefore cannot be applied when data is missing. On the other hand, our work focused both on extraction of the scores, when documented (stage 1) and also on developing a model to predict the scores (stage 2) and evaluate these against a gold standard. Our model can therefore be used to predict NIHSS from notes, even when missing or not documented.

Other studies have predicted stroke severity by approaching the data as a binary task problem [32,33], and thus are not directly comparable to ours. In one study [33], a 3D-Convolutional Neural Network was trained to predict low (NIHSS < 5) vs. high (NIHSS ≥ 5) based on preprocessed diffusion-weighted imaging (DWI) images. The NIHSS category was predicted at admission and on day 7 of hospitalization achieving an area under the receiver-operating characteristic curve (AUC) of 0.85 and 0.90, respectively. However, limiting to a binary classification limits the utility of the model, especially in light of a NIHSS of 6 being put in the same category as an NIHSS of 42. In a different study [32], the authors developed a random forests model that achieved a recall, precision and F1-score of 0.91 to predict improvement vs. worsening of NIHSS progression as a binary outcome from hospital admission until discharge, using baseline data within the first 72 hours of admission. By predicting NIHSS as a continuous variable, and enabling the prediction of NIHSS even when it is not documented or is missing from the EHR, our model overcomes the limitations of these prior works.

Our study has some limitations. While we validated our model in data from a different hospital, both hospitals are located in the same geographic region (Boston, Massachusetts). Nevertheless, the hospitals have different EHR systems, providers, and note types that increase generalizability. Future studies are required to assure generalizability of the model in other US and non-US populations. In future studies we will utilize the model across different hospitals and EHR systems in the US. Our model performance decreased for NIHSS scores greater than 30 due to fewer cases with high scores. Therefore, the model was not able to fully learn the patterns or main drivers for prediction of scores in that range. However, on closer examination of these cases, the patients’ clinical exams were very poor, with fewer clinical details documented for motor exam. Other cases included patients with fluctuating symptoms in the notes, making it difficult for the model to provide an accurate prediction. Future studies should investigate the model performance when developing the model with cohorts including higher NIHSS scores, using the methodology here presented. Finally, the model was trained with data from admission notes, and it was externally validated in discharge summary notes. Nonetheless, we did assess performance in a hold-out test set of admission notes and observed that the error was approximately the same. Additionally, the discharge summaries included the initial history and physical along with the admission exam. Another limitation consisted of the relatively small sample size of the MIMIC cohort, with a different distribution of NIHSS scores, compared to that of the MGH cohort. Thus, in future work we aim to validate the model in other cohorts with more diverse score distributions.

The automatic NLP model presented herein enables automatic retrieval of NIHSS scores from unstructured data, thereby enabling large-scale stroke severity phenotyping from electronic health records. This work overcomes key limitations of prior models that use administrative data or NLP for prediction of stroke severity. Our model can enable real world research and quality improvement studies to address process improvement, outcomes research, and health disparities in stroke care. Future directions include validating this model in additional electronic health datasets.

## Data Availability

The clinical de-identified data used in this manuscript will be publicly available in a designated repository, upon acceptance of the manuscript.

## Non-standard Abbreviations and Acronyms

AUC: Area under the receiver-operating characteristic curve
BIDMC: Beth Israel Deaconess Medical Center
CI: Confidence intervals
CONSORT: Consolidated Standards of Reporting Trials
CPT: Current Procedural Terminology
EHRs: Electronic health records
ICD: International Classification of Diseases
ICU: Intensive care unit
LASSO: Least absolute shrinkage and selection operator
MCA: Middle cerebral artery
MGH: Massachusetts General Hospital
MIMIC-III: Medical Information Mart for Intensive Care III
NIHSS: National Institutes of Health Stroke Scale
NLP: Natural language processing
RMSE: Root mean squared error
SC: Spearman correlation
STROBE: STrengthening the Reporting of OBservational studies in Epidemiology

## Acknowledgements

This work was funded by NIH R01NS131347 SFZ.

## Sources of Funding

Dr. Westover was supported by grants from the NIH (RF1AG064312, RF1NS120947, R01AG073410, R01HL161253, R01NS126282, R01AG073598, R01NS131347, R01NS130119), and NSF (2014431). Dr. Sahar F. Zafar was supported by the NIH (K23NS114201, R01NS126282, R01AG082693).

## Disclosures

Dr. Zafar is a clinical neurophysiologist for Corticare, received speaking honoraria from Marinus, and received royalties from Springer publishing, unrelated to this work. Dr. Westover is a co-founder, scientific advisor, and consultant to Beacon Biosignals and has a personal equity interest in the company. He receives royalties for authoring Pocket Neurology from Wolters Kluwer and Atlas of Intensive Care Quantitative EEG by Demos Medical. None of these interests played any role in the present work.

## Notes

### Competing Interest Statement

The authors have declared no competing interest.

### Clinical Trial

The study consists of a retrospective data analysis and it was approved by the Mass General Brigham Institutional Review Board; a waiver of informed consent was obtained for this observational study.

## References

1. Fonarow GC, Kapral MK, Schwamm LH. Future of Quality and Outcomes Research in Stroke. Circ Cardiovasc Qual Outcomes. 2015 Oct;8(6_suppl_3):S66–8.

2. Payne TH. The electronic health record as a catalyst for quality improvement in patient care. Heart Br Card Soc. 2016 Nov 15;102(22):1782–7.

3. Campanella P, Lovato E, Marone C, Fallacara L, Mancuso A, Ricciardi W, Specchia ML. The impact of electronic health records on healthcare quality: a systematic review and meta-analysis. Eur J Public Health. 2016 Feb;26(1):60–4.

4. Smith MA, Gigot M, Harburn A, Bednarz L, Curtis K, Mathew J, Farrar-Edwards D. Insights into measuring health disparities using electronic health records from a statewide network of health systems: A case study. J Clin Transl Sci. 2023;7(1):e54.

5. Rumball-Smith J, Bates DW. The Electronic Health Record and Health IT to Decrease Racial/Ethnic Disparities in Care. J Health Care Poor Underserved. 2018;29(1):58–62.

6. Adane K, Gizachew M, Kendie S. The role of medical data in efficient patient care delivery: a review. Risk Manag Healthc Policy. 2019 Apr 24;12:67–73.

7. 1. Uslu A, Stausberg J. Value of the Electronic Medical Record for Hospital Care: Update From the Literature. J Med Internet Res. 2021 Dec 23;23(12):e26323.

8. Wani D, Malhotra M. Does the meaningful use of electronic health records improve patient outcomes? J Oper Manag. 2018 May 1;60:1–18.

9. Lyden P. Using the National Institutes of Health Stroke Scale. Stroke. 2017 Feb;48(2):513– 9.

10. Beaulieu-Jones BK, Lavage DR, Snyder JW, Moore JH, Pendergrass SA, Bauer CR. Characterizing and Managing Missing Structured Data in Electronic Health Records: Data Analysis. JMIR Med Inform. 2018 Feb 23;6(1):e11.

11. PCORI-National-Priorities-and-Research-Agenda-2012-05-21-FINAL1.pdf [Internet]. [cited 2024 Feb 6]. Available from: https://www.pcori.org/assets/PCORI-National-Priorities-and-Research-Agenda-2012-05-21-FINAL1.pdf

12. Institute of Medicine. Initial National Priorities for Comparative Effectiveness Research [Internet]. Washington, D.C.: National Academies Press; 2009 [cited 2024 Feb 6]. Available from: http://www.nap.edu/catalog/12648

13. Leifer D, Bravata DM, Connors JJ (Buddy), Hinchey JA, Jauch EC, Johnston SC, Latchaw R, Likosky W, Ogilvy C, Qureshi AI, et al. Metrics for Measuring Quality of Care in Comprehensive Stroke Centers: Detailed Follow-Up to Brain Attack Coalition Comprehensive Stroke Center Recommendations. Stroke. 2011 Mar;42(3):849–77.

14. Jovin TG, Albers GW, Liebeskind DS, STAIR IX Consortium. Stroke Treatment Academic Industry Roundtable: The Next Generation of Endovascular Trials. Stroke. 2016 Oct;47(10):2656–65.

15. Williams LS, Yilmaz EY, Lopez-Yunez AM. Retrospective assessment of initial stroke severity with the NIH Stroke Scale. Stroke. 2000 Apr;31(4):858–62.

16. Kasner SE, Chalela JA, Luciano JM, Cucchiara BL, Raps EC, McGarvey ML, Conroy MB, Localio AR. Reliability and validity of estimating the NIH stroke scale score from medical records. Stroke. 1999 Aug;30(8):1534–7.

17. Song S, Fonarow GC, Olson DM, Liang L, Schulte PJ, Hernandez AF, Peterson ED, Reeves MJ, Smith EE, Schwamm LH, Saver JL. Association of Get With The Guidelines-Stroke Program Participation and Clinical Outcomes for Medicare Beneficiaries With Ischemic Stroke. Stroke. 2016 May;47(5):1294–302.

18. Specogna AV, Patten SB, Turin TC, Hill MD. The Reliability and Sensitivity of the National Institutes of Health Stroke Scale for Spontaneous Intracerebral Hemorrhage in an Uncontrolled Setting. PLOS ONE. 2013 Dec 19;8(12):e84702.

19. You S, Zheng D, Yoshimura S, Ouyang M, Han Q, Wang X, Cao Y, Delcourt C, Song L, Arima H et al. Optimum Baseline Clinical Severity Scale Cut Points for Prognosticating Intracerebral Hemorrhage: INTERACT Studies. Stroke. 2024 Jan;55(1):139–45.

20. Finocchi C, Balestrino M, Malfatto L, Mancardi G, Serrati C, Gandolfo C. National Institutes of Health Stroke Scale in patients with primary intracerebral hemorrhage. Neurol Sci Off J Ital Neurol Soc Ital Soc Clin Neurophysiol. 2018 Oct;39(10):1751–5.

21. 1. Andersen KK, Olsen TS, Dehlendorff C, Kammersgaard LP. Hemorrhagic and Ischemic Strokes Compared. Stroke. 2009 Jun;40(6):2068–72.

22. Kasner SE, Cucchiara BL, McGarvey ML, Luciano JM, Liebeskind DS, Chalela JA. Modified National Institutes of Health Stroke Scale Can Be Estimated From Medical Records. Stroke. 2003 Feb;34(2):568–70.

23. Kogan E, Twyman K, Heap J, Milentijevic D, Lin JH, Alberts M. Assessing stroke severity using electronic health record data: a machine learning approach. BMC Med Inform Decis Mak. 2020 Jan 8;20(1):8.

24. Yang L, Huang X, Wang J, Yang X, Ding L, Li Z, Li J. Identifying stroke-related quantified evidence from electronic health records in real-world studies. Artif Intell Med. 2023 Jun;140:102552.

25. Katzan IL, Spertus J, Bettger JP, Bravata DM, Reeves MJ, Smith EE, Bushnell C, Higashida RT, Hinchey JA, Holloway RG, et al. Risk adjustment of ischemic stroke outcomes for comparing hospital performance: a statement for healthcare professionals from the American Heart Association/American Stroke Association. Stroke. 2014 Mar;45(3):918– 44.

26. Fonarow GC, Alberts MJ, Broderick JP, Jauch EC, Kleindorfer DO, Saver JL, Solis P, Suter R, Schwamm LH. Stroke outcomes measures must be appropriately risk adjusted to ensure quality care of patients: a presidential advisory from the American Heart Association/American Stroke Association. Stroke. 2014 May;45(5):1589–601.

27. Reeves MJ, Smith EE, Fonarow GC, Zhao X, Thompson M, Peterson ED, Schwamm LH, Olson D. Variation and Trends in the Documentation of National Institutes of Health Stroke Scale in GWTG-Stroke Hospitals. Circ Cardiovasc Qual Outcomes. 2015 Oct;8(6 Suppl 3):S90–98.

28. Vandenbroucke JP, Elm E von, Altman DG, Gøtzsche PC, Mulrow CD, Pocock SJ, Poole C, Schlesselman JJ, Egger M. Strengthening the Reporting of Observational Studies in Epidemiology (STROBE): Explanation and Elaboration. PLOS Med. 2007 out;4(10):e297.

29. Johnson AE, Pollard TJ, Shen L, Lehman L wei H, Feng M, Ghassemi M, Moody B, Szolovits P, Celi LA, Mark RG. MIMIC-III, a freely accessible critical care database. Sci Data. 2016 May 24;3(1):160035.

30. Mayampurath A, Parnianpour Z, Richards CT, Meurer WJ, Lee J, Ankenman B, Perry O, Mendelson SJ, Holl JL, Prabhakaran S. Improving Prehospital Stroke Diagnosis Using Natural Language Processing of Paramedic Reports. Stroke. 2021 Aug;52(8):2676–9.

31. Guan W, Ko D, Khurshid S, Lipsanopoulos ATT, Ashburner JM, Harrington LX, Rost NS, Atlas SJ, Singer DE, McManus DD, Anderson CD, Lubitz SA. Automated Electronic Phenotyping of Cardioembolic Stroke. Stroke. 2021 Jan;52(1):181–9.

32. Gkantzios A, Kokkotis C, Tsiptsios D, Moustakidis S, Gkartzonika E, Avramidis T, Tripsianis G, Iliopoulos I, Aggelousis N, Vadikolias K. From Admission to Discharge: Predicting National Institutes of Health Stroke Scale Progression in Stroke Patients Using Biomarkers and Explainable Machine Learning. J Pers Med. 2023 Sep 14;13(9):1375.

33. Zeng Y, Long C, Zhao W, Liu J. Predicting the Severity of Neurological Impairment Caused by Ischemic Stroke Using Deep Learning Based on Diffusion-Weighted Images. J Clin Med. 2022 Jul 11;11(14):4008.

## References in Supplementary material

1. Johnson AE, Pollard TJ, Shen L, Lehman L wei H, Feng M, Ghassemi M, Moody B, Szolovits P, Celi LA, Mark RG. MIMIC-III, a freely accessible critical care database. Sci Data. 2016 May 24;3(1):160035.

2. Yang L, Huang X, Wang J, Yang X, Ding L, Li Z, Li J. Identifying stroke-related quantified evidence from electronic health records in real-world studies. Artif Intell Med. 2023 Jun;140:102552.

3. Woodfield R, Grant I, Group UBSO, Group UBFU and OW, Sudlow CLM. Accuracy of Electronic Health Record Data for Identifying Stroke Cases in Large-Scale Epidemiological Studies: A Systematic Review from the UK Biobank Stroke Outcomes Group. PLOS ONE. 2015 Oct 23;10(10):e0140533.

4. Mitchell JD, Collen JF, Petteys S, Holley AB. A simple reminder system improves venous thromboembolism prophylaxis rates and reduces thrombotic events for hospitalized patients1. J Thromb Haemost JTH. 2012 Feb;10(2):236–43.

5. Reeves MJ, Smith EE, Fonarow GC, Zhao X, Thompson M, Peterson ED, Schwamm LH, Olson D. V. Variation and Trends in the Documentation of National Institutes of Health Stroke Scale in GWTG-Stroke Hospitals. Circ Cardiovasc Qual Outcomes. 2015 Oct;8(6 Suppl 3):S90–98.

6. Porter MF. An algorithm for suffix stripping. Program Electron Libr Inf Syst [Internet]. 1980 Jan 1 [cited 2023 Dec 27]; Available from: https://www.scienceopen.com/document?vid=49b876ff-1ee1-447f-9eb7-95a1d69d999b

7. Tibshirani R. Regression Shrinkage and Selection via the Lasso. J R Stat Soc Ser B Methodol. 1996;58(1):267–88.

